# The prevalence and topography of spinal cord demyelination in multiple sclerosis: a retrospective study

**DOI:** 10.1101/2022.06.14.22276413

**Authors:** Alex D. Waldman, Cecilia Catania, Marco Pisa, Mark Jenkinson, Michael J. Lenardo, Gabriele C. DeLuca

## Abstract

Spinal cord pathology is a major determinant of irreversible disability in progressive multiple sclerosis. The demyelinated lesion is a cardinal feature. The well-characterised anatomy of the spinal cord and new analytic approaches allows the systematic study of lesion topography and its extent of inflammatory activity unveiling new insights into disease pathogenesis. We studied cervical, thoracic, and lumbar spinal cord tissue from 119 pathologically confirmed multiple sclerosis cases. Immunohistochemistry was used to detect demyelination (PLP) and classify lesional inflammatory activity (CD68). Prevalence and distribution of demyelination, staged by lesion activity, was determined and topographical maps were created to identify patterns of lesion prevalence and distrtibution using mixed models and permutation-based voxelwise analysis. 460 lesions were observed throughout the spinal cord with 76.5% of cases demonstrating at least 1 lesion. The cervical level was preferentially affected by lesions. 58.3% of lesions were inflammatory with 87.9% of cases harbouring at least 1 inflammatory lesion. Topographically, lesions consistently affected the dorsal and lateral columns with relative sparing of subpial areas in a distribution mirroring the vascular network. The presence of spinal cord lesions and the proportion of active lesions related strongly with clinical disease milestones, including time from onset to wheelchair and onset to death. We demonstrate that spinal cord demyelination is common, highly inflammatory, has a predilection for the cervical level, and relates to clinical disability. The topography of lesions in the dorsal and lateral columns and relative sparing of subpial areas points to a role of the vasculature in lesion pathogenesis, suggesting short-range cell infiltration from the blood and signaling molecules circulating in the perivascular space incite lesion development. These findings challenge the notion that end-stage progressive multiple sclerosis is ‘burnt out’ and an outside-in lesional gradient predominates in the spinal cord. Taken together, this study provides support for long-term targeting of inflammatory demyelination in the spinal cord and nominates vascular dysfunction as a potential target for new therapeutic approaches to limit irreversible disability.

## Introduction

The destruction of the myelin covering nerve axons, i.e. demyelination, is the central hallmark of MS pathology [10, 13, 16, 17, 38, 43]. While lesions were originally thought to be restricted to the white matter, early studies revealed that grey matter was not spared [6, 20, 38]. Heterogeneity in the extent and distribution of demyelination along the length of the neuraxis is a major contributor to the accumulation of irreversible disability [26, 37]. The spinal cord is a key site where demyelination causes the predominant sensorimotor deficits of the disease, especially during progression [39].

The aetiopathogenesis of demyelinated lesions in MS is still debated. Early observations showed that the topography of spinal cord lesions mirrors the architecture of the venous system [2, 13, 15, 16]. Also, lesions are commonly found in the upper cervical cord in close opposition to the denticulate ligaments where the tensile forces on the vasculature are likely strongest [32]. In contrast to this vascular-centric view of spinal cord lesion pathogenesis, a more recent MRI study has observed an ‘outside-in’ gradient of demyelination in the cervical spinal cord that is most pronounced at surfaces exposed to cerebrospinal fluid (CSF) [33].

Given the lack of consensus regarding spinal cord demyelination, we undertook the largest neuropathological study of staged lesion topography along the length of the spinal cord. Compared to MRI, neuropathology is indisputedly more sensitive and specific for lesion evaluation and provides detailed information on the extent and nature of inflammation in the MS spinal cord which we compared to longitudinal clinical measures.

We show that spinal cord demyelination is frequent, highly inflammatory, and correlates with rapid disability accumulation in our series of MS cases. Analyses of lesion topography show a dorsal and lateral column distribution with relative sparing of the subpial circumferential area. Notably, we did not observe an ‘outside-in’ gradient of lesions but rather most lesions were distant from the CSF interface. We conclude that preventing inflammatory demyelination even at the late stages of progressive disease may be a useful therapeutic approach to limit irreversible disability and further investigation of the role of the vasculature in this process is warranted.

## Methods

### Study Population

A post-mortem cohort (n = 119) of pathologically-confirmed MS cases derived from the UK MS Tissue Bank was obtained for study. Cases were selected based on the availability of formalin-fixed-frozen, paraffin-processed cervical, thoracic, and lumbar spinal cord material. Demographic, clinical, and pathological information were extracted from post-mortem reports (**Table 1**). A subset of these cases (n = 68) also had motor cortical blocks, sampled from mesial precentral gyrus, available for comparative analyses. Ethical approval was obtained (REC# 08/MRE09/31+5) and Human Tissue Act Guidelines were followed.

**Table 1.** Cohort Demographics.

|  |  | **Multiple Sclerosis (n=119)** |
| --- | --- | --- |
| **Number of Samples** | *Cervical* | *102* |
|  | *Thoracic* | *115* |
|  | *Lumbar* | *106* |
| **Disease Type\*** | *Primary Progressive* | 14 |
|  | *Secondary Progressive* | 100 |
|  | *Progressive-Relapsing* | 4 |
|  | *Relapsing-Remitting* | 1 |
| **Age (Mean ± SD)** |  | 61.7 ± 12.5 |
| **Sex (M:F)** |  | 36:83 |
| **Disease Duration in Years (Mean ± SD)\*** |  | 28.4 ± 11.2 |
| **Time to Wheelchair (Mean ± SD)\*** |  | 17.3 ± 10.3 |
| **PMI in Hours (Mean ± SD)** |  | 16.7 ± 6.6 |

SD=standard deviation, PMI=post-mortem interval
\*Data missing for disease duration (n=6) and time to wheelchair (n=20)

### Spinal Cord Neuropathological Assessment

Characterisation of lesion distribution and inflammatory activity was performed using the workflow described in **Fig. 1**. Formalin-fixed-frozen, paraffin-processed material from the cervical, thoracic, and lumbar spinal cord was serially sectioned (6 μm thick) before immunohistochemical studies to label myelin and macrophage inflammation by the proteolipid protein (PLP) and CD68 primary antibodies, respectively. Immunohistochemical optimisation determined the appropriate antigen retrieval method, primary antibody concentration, and incubation conditions for staining (**Supplementary Table 1, online resource**). In short, tissue was first deparaffinised and rehydrated. Endogenous peroxidase activity was quenched with 3% hydrogen peroxide prior to heat-induced antigen retrieval. Staining was performed using the semiautomatic Shandon Sequenza system. The Dako REAL™ EnVision™ Detection System was used for 3,3’-diaminobenzidine (DAB) detection. Sections were counter-stained with hematoxylin. Omission of primary antibodies confirmed the specificity of the immunoreactions.

**Fig. 1.**
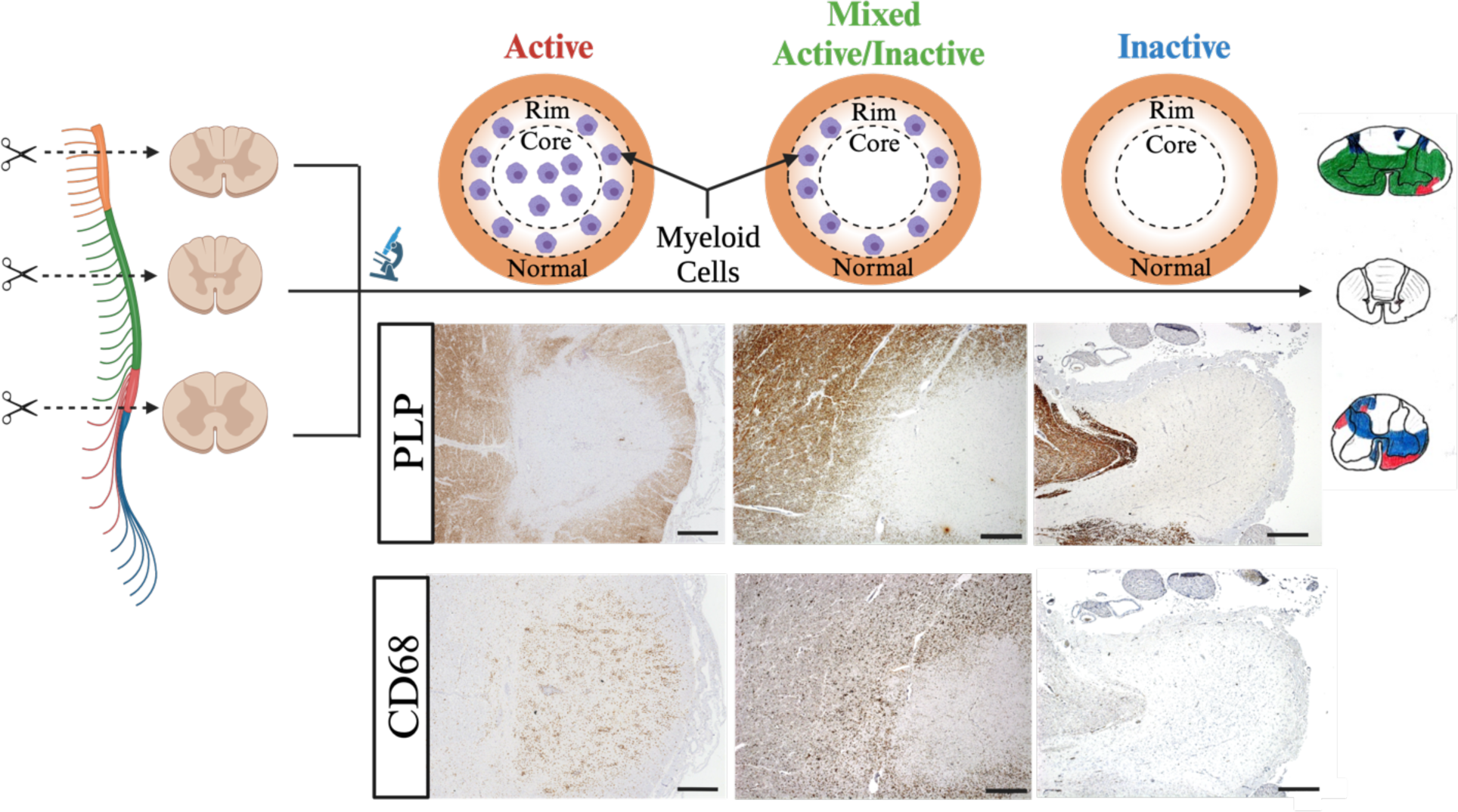
Methodological Approach to Assessing Staged Lesion Topography. Spinal cord material from the cervical, thoracic, and lumbar region was sectioned and processed for immunohistochemistry to stain for myelin (PLP) and activated macrophages (CD68). Light microscopy was used to identify areas of complete myelin absence and infiltration by myeloid cells was used to stage each lesion (scale bars: 0.5mm). Staged lesions were carefully drawn onto standardized templates prior to downstream computational analyses.

Slides were viewed on an Olympus BX43 Brightfield Microscope. Areas of demyelination were defined by complete loss of myelin in PLP-stained sections. Inflammatory activity of demyelinated lesions was assessed using established criteria [28]. Lesion activity was classified broadly as 1) active, 2) mixed active/inactive, and 3) inactive by histologically assessing the abundance and distribution of myeloid cells in CD68-stained sections. Specifically, active lesions were defined by the presence of homogenous CD68+ hypercellularity throughout the lesion, mixed active/inactive lesions by a hypocellular lesion core and rim of CD68+ cells along the border, and inactive lesions by a relative paucity of CD68+ cells throughout the lesion. Lesions classified as active, mixed active/inactive, and inactive were drawn by consensus between a trained microscopist (CC) and experimental neuropathologist (GCD) onto standardised templates of cervical, thoracic, and lumbar spinal cord levels ensuring that proportional lesion location and size were respected. For a subset of analyses, active and mixed/active inactive lesion categories were combined into a single category of ‘inflammatory lesions’ and compared to inactive lesions. Representative histology images were captured using a Zeiss AxioCam MRc Color Camera. Hand-drawn lesion maps were digitised and then analysed using the FMRIB Software Library (FSL) for downstream analyses [24].

### Motor Cortical Neuropathological Assessment

Formalin-fixed-frozen, paraffin-processed material from the motor cortex was serially sectioned (6 μm thick) before immunohistochemical studies to label myelin with proteolipid protein (PLP). Antibody conditions and immunohistochemical procedure were identical to those use in the spinal cord (**Supplementary Table 1, online resource**).

Slides were scanned using the Aperio Scanscope AT Turbo system (Leica Biosystems) prior to viewing in QuPath v0.4.3. Cortical lesions were identified as described previously [45]. In short, as with the assessment of spinal cord material, lesions were defined as areas of complete absence of PLP immunolabelling. Given that the cortical ribbon was commonly incomplete, areas of demyelination were only notated in regions where all 6 cortical layers were present. Cases were then dichotomised into those that exhibited subpial demyelination (n = 38) and those that did not (n = 30) for downstream analyses.

### Statistical Analysis

Graphical representations of all results were created in Prism 10 and BioRender. Statistical analysis was performed in RStudio Version 2022.7.1.554 running R 4.2.1. Both case-level and lesion-level analyses were performed. For case-level analyses, lesion presence was used as an input for mixed effects logistic regression. Models included fixed effects terms of spinal cord level, lesion stage, and location (anterior, posterior, and lateral funiculi), where relevant, with a random effect of case. When including lesion stage as a fixed effect, inflammatory and inactive were treated as mutually exclusive categories. The presence of at least 1 inflammatory lesion defined a case or spinal cord level as inflammatory whilst the remaining were defined as inactive. In an additional analysis, lesion stage was modelled directly as a dependent variable using mixed effects logistic regression. For lesion-level analyses, lesion counts were used as an input for mixed effects poisson regression. An offset term representing the total number of lesions identified per-case was used to allow for modelling of lesion proportions rather than counts. Models included fixed effects terms of spinal cord level, lesion stage, and location (anterior, posterior, and lateral funiculi), where relevant, with a random effect of case. In these analyses, lesion stage was defined with either a two-category (inflammatory and inactive) or three-category (active, mixed/active inactive, and inactive) schema. All modelling above was performed using the glmmTMB package. The proportion of the spinal cord circumference impacted by subpial demyelination was used as the dependent variable for mixed effects quasi-binomial regression fit using the MASS package [42]. Models included fixed effects terms of spinal cord level and lesion stage, and the presence of motor cortical subpial disease, where relevant, with a random effect of case. Sex and age were entered as covariates when the presence of motor cortical subpial disease was the between-subjects fixed effect of interest. The emmeans package was used to extract estimated marginal means and assess the significance of pairwise comparisons [40]. A bias adjustment based on the value of sigma (standard deviation of the random effect) was applied. P-values were adjusted using the multivariate adjustment method. An adjusted p-value of 0.05 was used to define statistically significant comparisons. The caret package was used to calculate the sensitivity and specificity of cervical lesion presence in determining the liklehood of lesion presence at other levels of the spinal cord. The survival package was used to fit multivariate cox proportional hazards models to understand the effects of lesion presence, inflammatory activity, subpial lesion presence, and staged lesion proportion on clinical milestones obtained during life (time from onset wheelchair and onset to death) whilst controlling for sex and age of onset [11].

FSL tools were used to generate lesion frequency heatmaps. The Randomise function was used to perform permutation-based voxel-wise analysis with 500 permutations to identify lesion predilection sites for lesions also taking into account inflammatory stage. Confounders (age and sex) were included in the model to remove any potential statistical abberations. Distance-based lesion segmentation was performed according to the method described in Ouellette *et al.* 2020. In short, 10 concentric distance zones (termed bins) were generated for the cervical, thoracic, and lumbar spinal cord template using the distancemap function in FSL, based on a the central canal. For each patient, the normalised lesion fraction (lesional area within each bin divided by the area of the bin) was calculated. A one-way ANOVA was used to test for differences in lesion area across bins for each level separately. Post-hoc tests with Dunnet’s correction were used to assess differences between all bins and bin 5 (the innermost bin furthest from the CSF).

## Results

### Demographics

The cohort of 119 progressive MS cases comprised 102 cervical, 116 thoracic, and 107 lumbar spinal cord sections. Cases with varied demographic profiles were included to maximise generalisability of the results (**Table 1**). 83 cases were female (69.7%) with a cohort mean age of 61.7 ± 12.5 years, disease duration of 28.4 ± 11.2 years, time from onset to wheelchair of 17.3 ± 10.3 years, and post-mortem interval of 16.7 ± 6.6 hours.

### Prevalence of Spinal Cord Demyelination and Inflammatory Activity

At least one spinal cord lesion was observed in 91/119 (76.5%) of cases with 80/91 (87.9%) harbouring an inflammatory lesion (**Fig. 2a**). Lesions were more likely to be present in the cervical compared to the lumbar region in a categorical variable analysis (OR = 10.01, SE = 3.96, p = 0.0056) with a significant linear trend observed in an ordinal variable analysis (Cervical > Thoracic > Lumbar, p=0.0018) (**Fig. 2b**). Inflammatory sections were more common than purely inactive sections at all spinal cord levels (Cervical: Probability = 0.84, SE = 0.065, p = 0.0001; Thoracic: Probability = 0.70, SE = 0.067, p = 0.0031; Lumbar: Probability = 0.73, SE = 0.074, p = 0.0022) though all spinal cord levels were equally likely to be inflammatory (**Fig. 2c**).

**Fig. 2.**
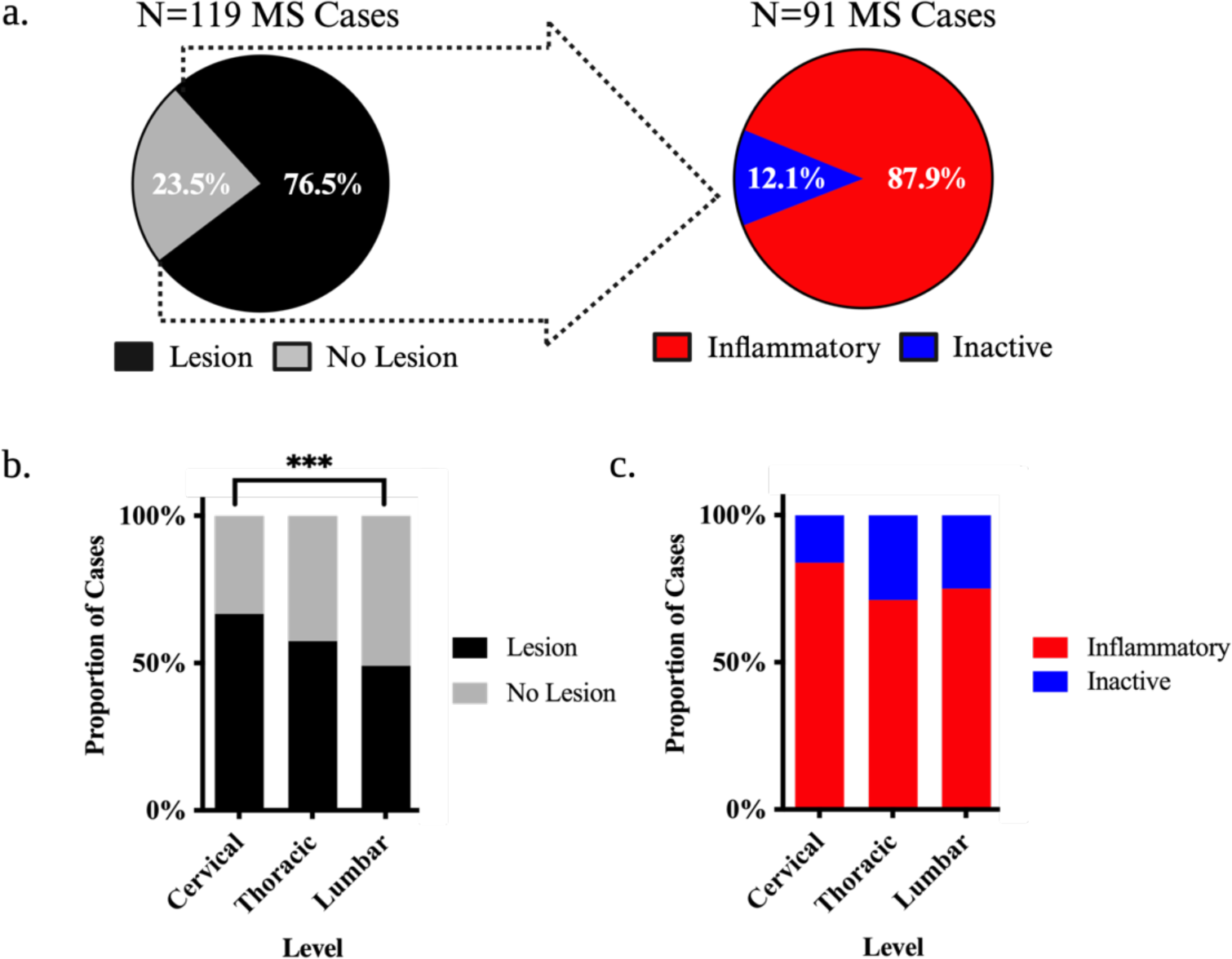
Case-Level Prevalence of Demyelination in the MS Spinal Cord. **(a)** Pie charts highlighting the observed proportion of cases with spinal cord lesions and those that harboured at least 1 inflammatory (active or mixed/active inactive) lesion. (**b-c**) Stacked bar charts depicting the proportion of MS cases that harboured lesions at each level of the spinal cord irrespective of stage (**b**) and classified by the presence of inflammation (**c**). Proportions represent observed values, and the asterisks indicate significant post-hoc pairwise comparisons following logistic mixed modelling and multivariate adjustment for multiple comparisons (*p<0.05; **p<0.01; ***p<0.001; ****p<0.0001).

In total, a total of 460 lesions were identified in the spinal cord of which 177 (38.5%) were cervical, 152 (33.0%) were thoracic, and 131 (28.5%) were lumbar. The proportion of lesions was higher in the cervical cord when compared to the lumbar region (Ratio = 1.36, SE = 0.156, p = 0.0216) with a significant linear trend (Cervical > Thoracic > Lumbar p = 0.0080) (**Fig 3a**). Across all cord levels, 268 lesions (58.3%) were inflammatory with 78 (17.0%) being active and 190 (41.3%) being mixed active/inactive. 192 lesions (41.7%) were inactive (**Fig 3b,d**). Overall, inflammatory lesions were more common than inactive lesions (Ratio = 0.72, SE = 0.068, p = 0.0004) (**Fig 3b**), a finding observed in cervical (Ratio = 0.65, SE = 0.10, p = 0.0058) and lumbar (Ratio = 0.66, SE = 0.12, p = 0.019) levels but not the thoracic level (Ratio = 0.85, SE = 0.14, p = 0.33) (**Fig 3c**). Mixed active/inactive and inactive lesions were more common than active lesions (Mixed active/inactive: Ratio = 2.46, SE = 0.331, p < 0.0001; Inactive: Ratio = 2.44, SE = 0.328, p < 0.0001) (**Fig 3d**), a finding that was observed at every cord level (Cervical: Active-Inactive Ratio = 2.50, SE = 0.56, p = 0.0002, Active-Mixed Active/Inactive Ratio = 2.82, SE = 0.62, p < 0.0001; Thoracic: Active-Inactive Ratio = 2.69, SE = 0.62, p < 0.0001, Active-Mixed Active/Inactive Ratio = 2.15, SE = 0.51, p = 0.0033; Lumbar: Active-Inactive Ratio = 2.17, SE = 0.54, p = 0.0050, Active-Mixed Active/Inactive Ratio = 2.29, SE = 0.56, p = 0.0019) (**Fig 3e**). Lesion proportions of all types were similar across levels (**Fig. 3c,e**).

**Fig. 3.**
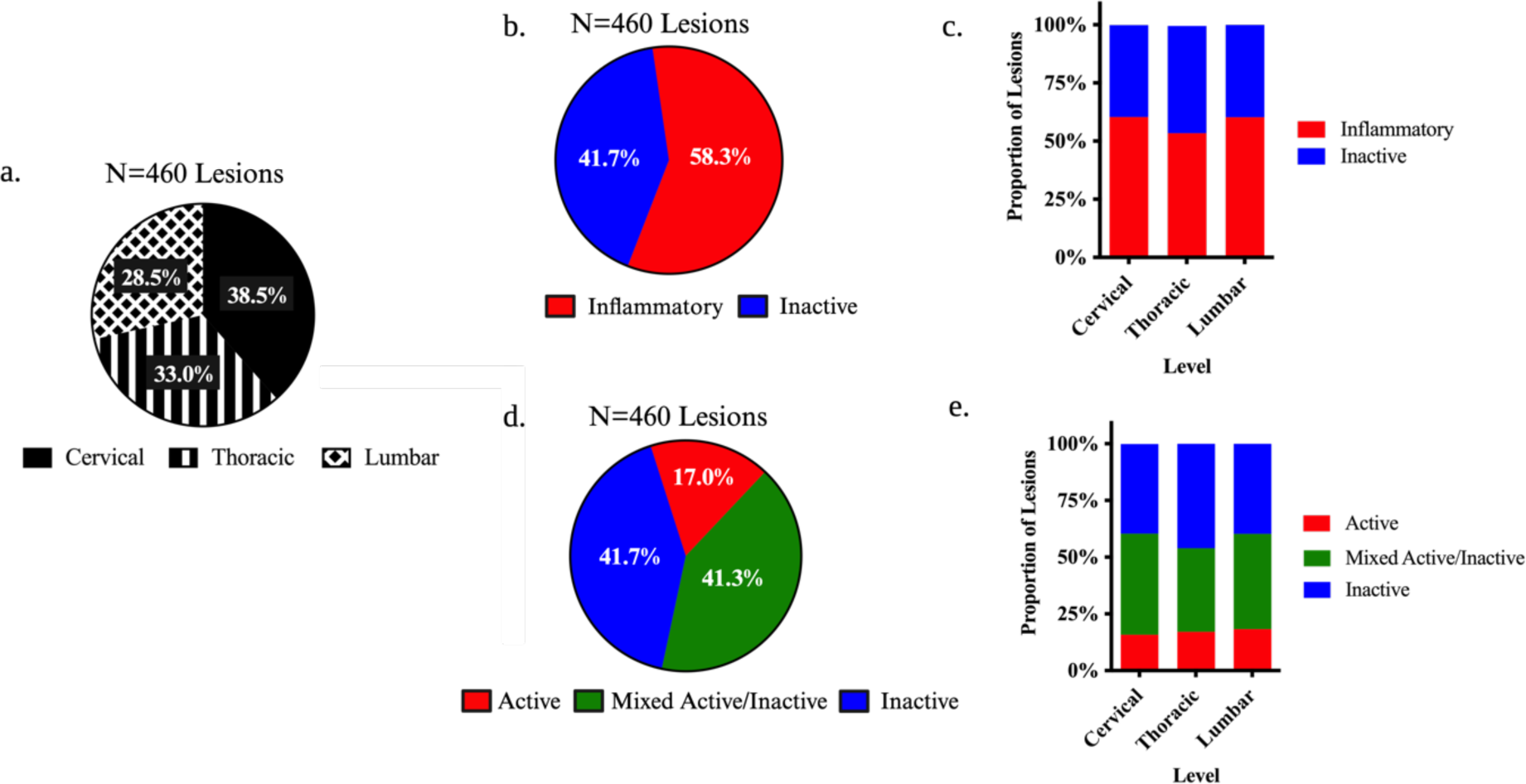
Lesion-Level Prevalence of Demyelination in the MS Spinal Cord. **(a)** Pie charts highlighting the proportion of lesions found within the cervical, thoracic, and lumbar spinal cord. (**b,d**) Pie charts depicting the proportion of lesions classified by the presence of inflammation across all levels. (**c,e**) Stacked bar chaerts illustrating the proportion of lesions at each cord level classified by inflammatory activity. Proportions represent observed values, and the asterisks indicate significant post-hoc pairwise comparisons following logistic mixed modelling and multivariate adjustment for multiple comparisons (*p<0.05; **p<0.01; ***p<0.001; ****p<0.0001).

### Concordance of Spinal Cord Lesion Presence Across Levels

We next explored whether the presence of a cervical spinal cord lesion impacted the likelihood of a lesion to be present in the thoracic and/or lumbar spinal cord. The sensitivity of a cervical spinal cord lesion to predict the presence of thoracic lesions was high (84.2%) though specificity was poor (59.5%), a finding also seen in the lumbar spinal cord (sensitivity = 86.0%; specificity =51.0%).

### Topography of Spinal Cord Demyelination and Inflammatory Activity

Representative histological patterns and heatmap summaries of lesion predilection sites demonstrated that the dorsal columns, lateral columns, and grey matter as a whole were consistently affected. Interestingly, the subpial surface was relatively spared at all spinal cord levels (**Fig. 4a-b**). A complementary analysis assessing the prevalence of anterior, posterior, and lateral funiculi involvement was undertaken (**Fig. 4c**). The probability of the lateral and posterior funiculi being impacted was consistently higher than the anterior funiculus (**Fig. 4d**).

**Fig. 4.**
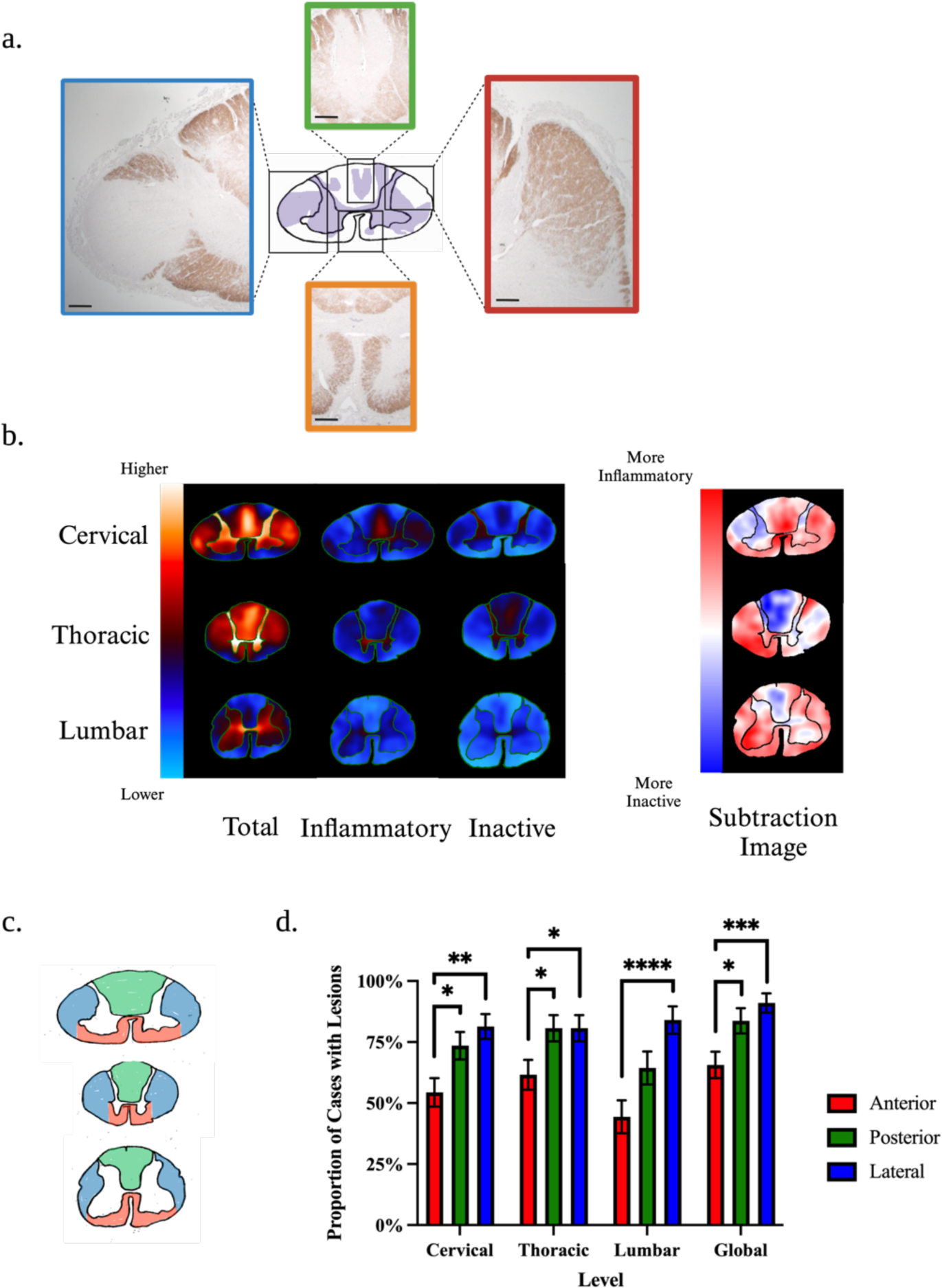
Pathological Patterns and Topography of Demyelination. **(a)** Representative histological images of lesion distribution patterns. The dorsal columns (green box), lateral columns (red box), and the whole grey matter (orange box) were consistently affected throughout the cohort (scale bars: 0.5mm). Interestingly, the subpial surface was commonly spared (red, green, and orange boxes). When present, subpial demyelination only affected a limited circumference of the cord (blue box). **(b)** Lesion frequency heatmaps of total, inflammatory, and inactive lesions in the cervical, thoracic, and lumbar spinal cord. (**c**) The subtraction images highlight areas more commonly affected by inflammatory lesions (red), inactive lesions (blue), and those equally affected by both types (white). **(c)** Anatomical landmarks of the anterior (red), lateral (blue) and posterior (green) funiculi. **(d)** Bar charts depicting the proportion of MS cases that exhibited demyelination of the anterior, posterior, and lateral funiculi at each level of the spinal cord and globally. Proportion values represent estimated marginal means and standard errors derived from mixed logistic regression models. Proportions add up to over 100% as cases can have involvement of more than one funiculus. The asterisks indicate significant post-hoc pairwise comparisons after the multivariate adjustment for multiple comparisons (*p<0.05; **p<0.01; ***p<0.001; ****p<0.0001).

### Prevalence and Extent of Demyelination at Cerebrospinal Fluid Interfaces

Given the relative subpial sparing, demyelination at CSF interfaces was analysed. 76/91 (83.5%) cases with lesions exhibited demyelination of the white matter abutting any part of the subpial surface of which 61/76 (80.3%) exhibited inflammatory demyelinating activity (**Fig. 5a**). Contrasts derived from logistic regression mixed models indicated that subpial involvement was not significantly more likely to be at any spinal cord level (**Fig. 5b**). Sections with inflammatory subpial involvement were more common than those found to be inactive at all levels with this relationship being significant at cervical and lumbar levels (Cervical: Probability = 0.72, SE = 0.0714, p = 0.0083; Thoracic: Probability = 0.64, SE = 0.071, p = 0.055; Lumbar: Probability = 0.69, SE = 0.074, p = 0.020) (**Fig. 5c**). When comparing across levels, sections from the cervical, thoracic, and lumbar spinal cord were equally likely to have inflammatory subpial disease (**Fig. 5c**).

**Fig. 5.**
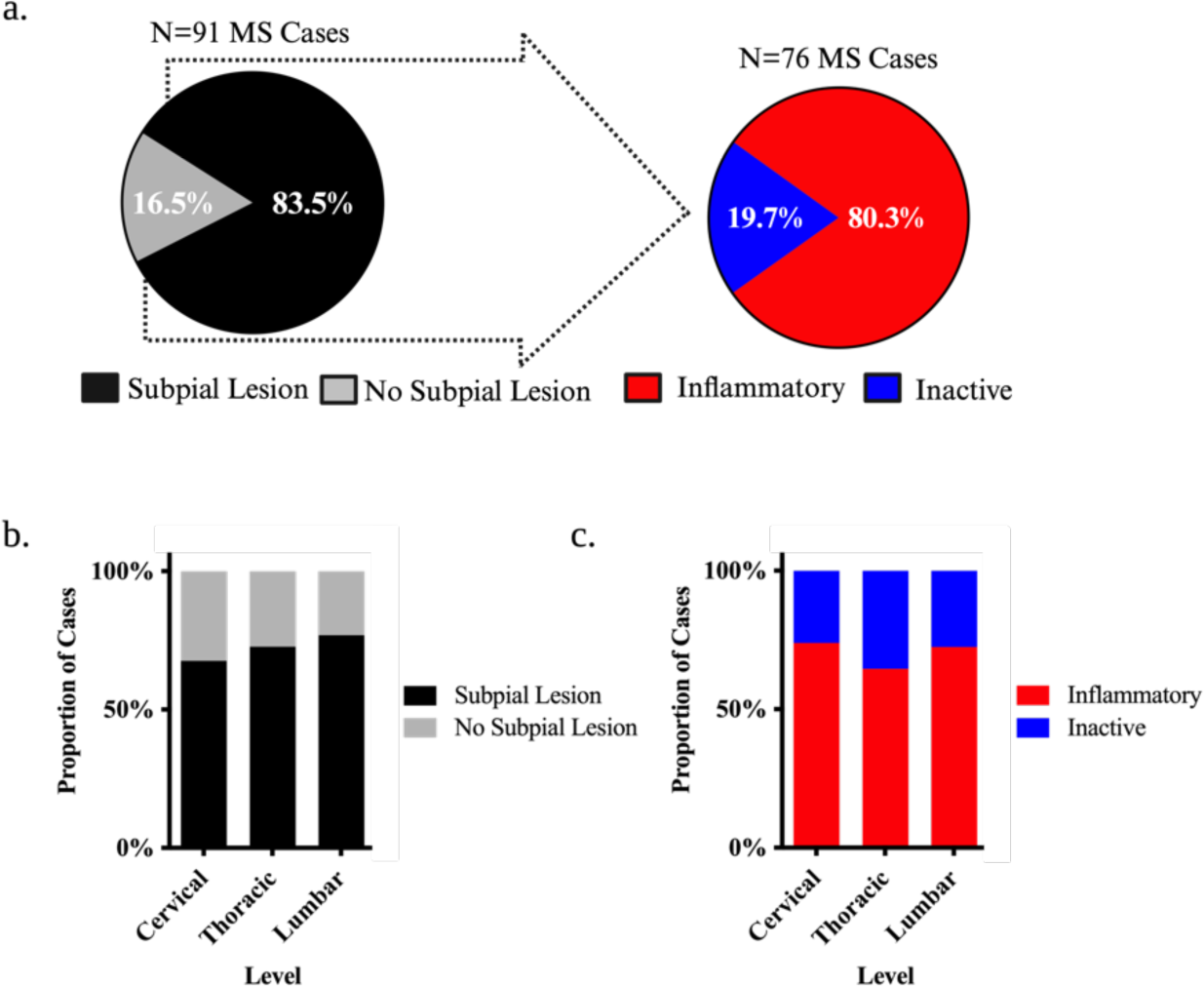
Prevalence of Subpial White Matter Demyelination in the MS Spinal Cord. **(a)** Pie charts highlighting the observed proportion of cases with subpial white matter spinal cord lesions and those that harboured at least 1 inflammatory (active or mixed/active inactive) subpial white matter lesion. (**b-c**) Stacked bar charts depicting the proportion of MS cases that harboured subpial white matter lesions at each level of the spinal cord irrespective of stage (**b**) and classified by the presence of inflammation (**c**). Proportions represent observed values, and the asterisks indicate significant post-hoc pairwise comparisons following logistic mixed modelling and multivariate adjustment for multiple comparisons (*p<0.05; **p<0.01; ***p<0.001; ****p<0.0001).

While presence of subpial demyelination was common across cases, the proportion of the subpial surface affected by lesions (i.e. spinal cord circumference affected by subpial lesions relative to total cord circumference) was lower than random chance (Cervical, Thoracic, and Lumbar: p < 0.001) and did not differ significantly across levels (Cervical: 19.2%, Thoracic: 23.2%, Lumbar: 19.7%) (**Fig. 6a**). When only considering cases with subpial lesions, the proportion of the subpial surface affected by lesions remained relatively low (Cervical: 29.3%, Thoracic: 32.5%, Lumbar: 26.4%) (**Fig. 6b**). When stratified by lesion stage, inflammatory lesions impacted a larger subpial circumference than inactive lesions in the cervical (Odds Ratio = 0.448, SE = 0.18, p = 0.049) and lumbar (Odds Ratio = 0.363, SE = 0.16, p = 0.025) spinal cord but not in the thoracic spinal cord (Odds Ratio = 1.20, SE = 0.47, p = 0.64) (**Fig 6c**). The proportional circumference impacted by active subpial lesions was significantly less than mixed active/inactive (Cervical: Odds Ratio = 24.83, p < 0.0001; Thoracic: Odds Ratio = 9.63, p = 0.0004; Lumbar: Odds Ratio = 6.70, p = 0.0012) and inactive lesions (Cervical: Odds Ratio = 13.49, p = 0.0007; Thoracic: Odds Ratio = 15.15, p < 0.0001; Lumbar: Odds Ratio = 4.10, p = 0.025) at all levels (**Fig. 6d**).

**Fig. 6.**
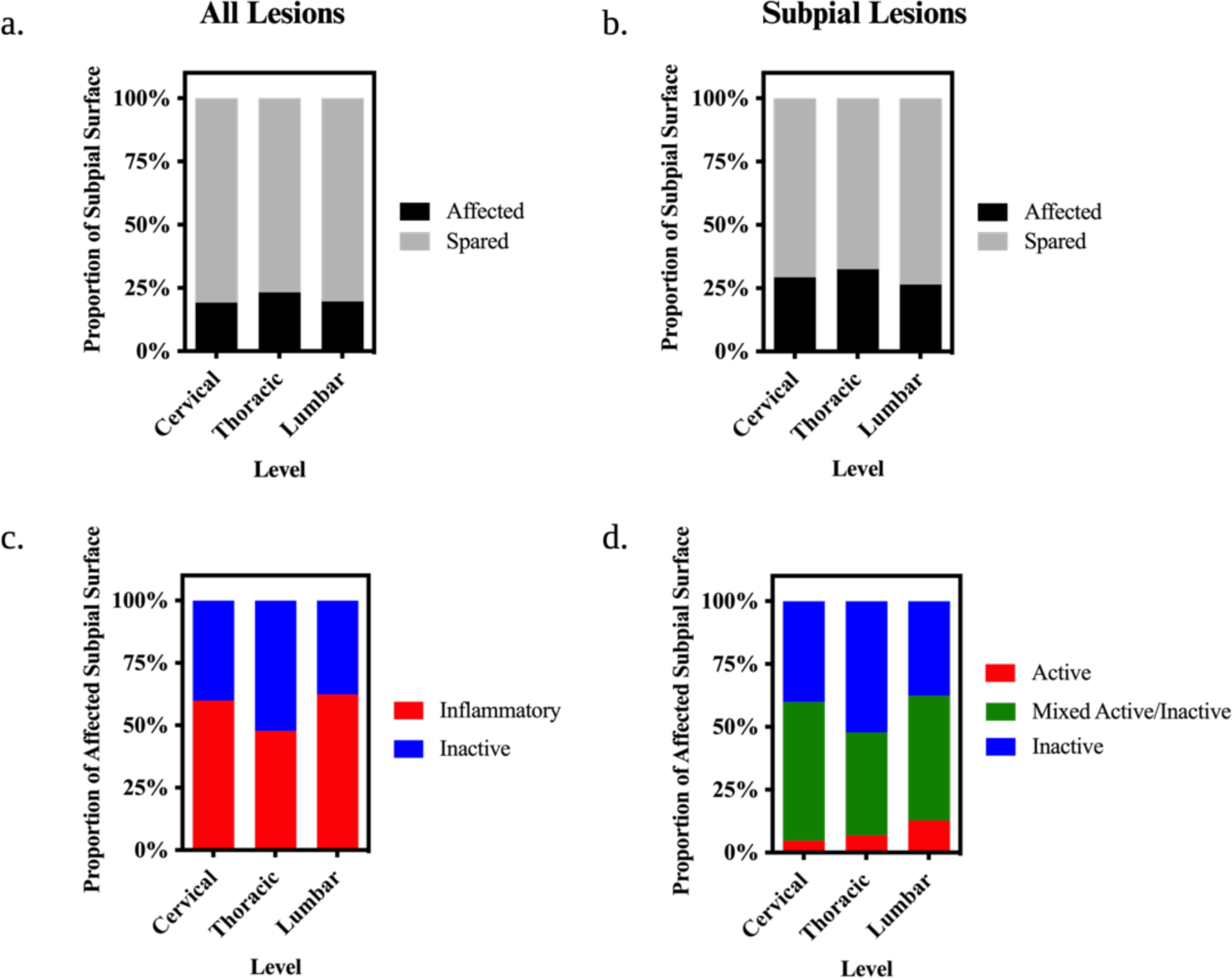
Extent of Subpial Demyelination in the MS Spinal Cord. **(a-b)** Stacked bar charts depicting the proportion of the subpial white matter surface affected by lesions at each level of the spinal cord irrespective of stage including cases with lesions of all types (**a**) and those with only subpial lesions (**b**). (c-d) Stacked bar charts depicting the proportion of the subpial white matter surface affected by lesions stratified by inflammatory activity. Proportions represent observed values, and the asterisks indicate significant post-hoc pairwise comparisons following linear mixed modelling and multivariate adjustment for multiple comparisons (*p<0.05; **p<0.01; ***p<0.001; ****p<0.0001).

We explored the relationship between subpial demyelination in the spinal cord and the well-documented pattern of subpial demyelination in the cerebral cortex [6, 21, 25]. 38/68 (55.9%) cases exhibited subpial demyelination in the motor cortex. Cases with evidence of cortical subpial demyelination did not demonstrate more extensive subpial demyelination at any level compared to cases with no subpial cortical demyelination (Cervical: Odds Ratio = 1.20, SE = 0.61, p = 0.73, Thoracic: Odds Ratio = 0.55, SE = 0.29, p = 0.079, Lumbar: Odds Ratio = 1.29, SE = 0.75, p = **0.86) (Supplementary Fig. 1, online resource).**

We also investigated the central canal, as it represents another CSF interface. 67/91 (73.6%) cases had lesions affecting the central canal with 46/67 (68.7%) categorised as inflammatory (**Supplementary Fig. 2a, online resource**). Contrasts derived from logistic regression mixed models indicated that central canal lesions were equally distributed at all spinal cord levels (**Supplementary Fig. 2b, online resource**). Sections with inflammatory central canal involvement were not more common than those that were found to be inactive at all levels. All spinal cord levels were also equally likely to have inflammatory disease of the central canal (**Supplementary Fig. 2c, online resource**).

Given that recent MRI work suggested a gradient of spinal cord lesions extending from fluid interfaces inwards (i.e. subpial or central canal to the parenchyma), we recapitulated the analysis by Ouellette *et al.* 2020. One-way ANOVA analysis demonstrated that normalised lesion volumes differed amongst bins at all levels (Cervical: F = 27.59, p < 0.0001; Thoracic: F=29.75, p < 0.0001; Lumbar: F = 19.85; p < 0.0001). Post-hoc analyses comparing all bins to the innermost bin (i.e. furthest from CSF boundaries) demonstrated a consistent decrease of normalised lesion volumes in the bins positioned furthest to the CSF (bin 1 and bin 10) (**Fig. 7a-c**).

**Fig. 7.**
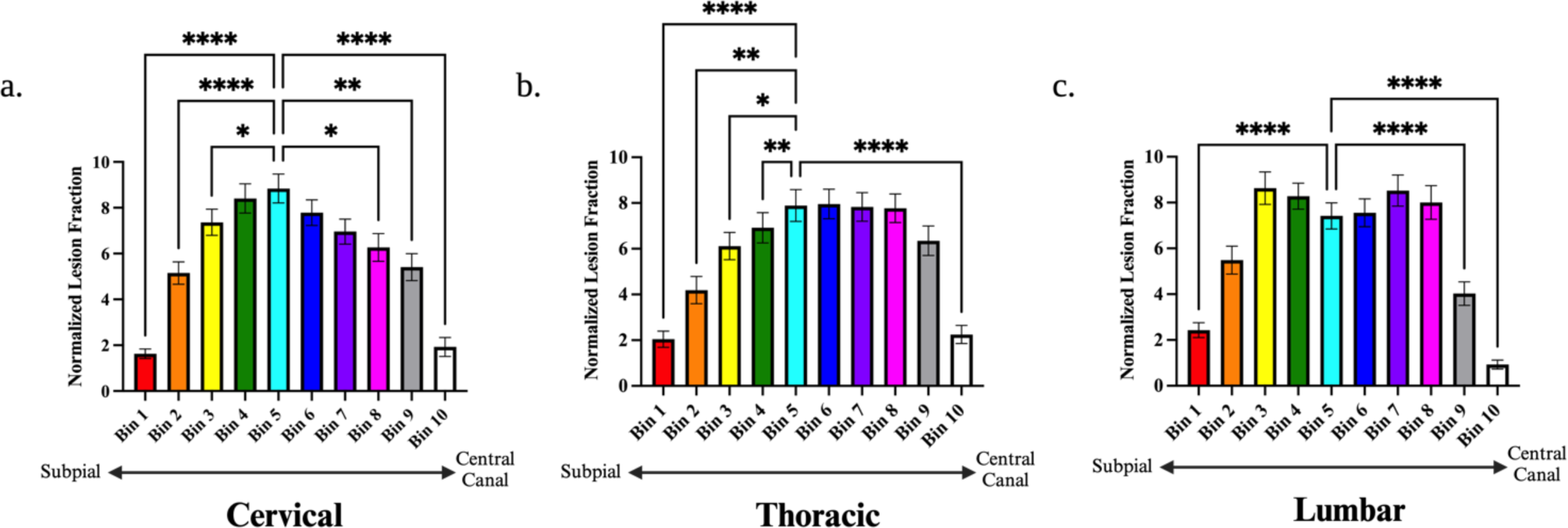
Distance-Based Lesion Segmentation Analysis of Spinal Cord Demyelination. Bar charts displaying normalised lesion fractions within concentric distance zones extending from the subpial surface (bin 1) to the central canal (bin 10) in the cervical (**a**), thoracic (**b**), and lumbar (**c**) spinal cord. Differences were assessed within each level individually with a one-way repeated measures ANOVA. Post-hoc comparisons with Dunnett’s correction were made compared to bin 5 (the region mid-way between the central canal and subpial surface). The asterisks indicate significant post-hoc pairwise comparisons (*p<0.05; **p<0.01; ***p<0.001; ****p<0.0001).

### Clinicopathological Relationships

Finally, we evaluated the relationship between spinal cord demyelination and clinical outcome. Time from disease onset to wheelchair and disease onset to death (disease duration) were used as surrogate markers of disease severity and response variables for multivariate cox proportional hazard analyses (**Fig. 8**). Sex and age of onset were included as covariates in all analyses. Cases with lesions anywhere in the spinal cord were more likely to be wheelchair-dependent (HR 2.45, 95% CI 1.43-4.20) or die (HR 2.65, 95% CI 1.61-4.34) at younger ages (**Fig. 8a-b**). Subpial location had no significant impact on clinical milestones (time to wheelchair: HR 1.32, 95% CI 0.71-2.44; time to death: (HR 1.71, 95% CI 0.93-3.13) (**Fig. 8c-d**). We also assessed the impact of inflammatory activity on clinical disease milestones. In a case-level analysis, cases with evidence of inflammatory activity did not exhibit a faster clinical decline (time to wheelchair: HR 1.01, 95% CI 0.53-1.93; time to death: HR 1.78, 95% CI 0.93-3.38) (**Fig. 8e-f**). However, a more granular analysis using lesion proportions demonstrated that a higher proportion of inflammatory lesions was observed in cases that died at younger ages (HR 2.76, 95% CI 1.45-5.29) though no relationship was observed with wheelchair trajectory (HR 1.01, 95% CI 0.53-1.94) (**Fig. 8g-h**). Interestingly, the relationship with time to death was driven by the proportion of active lesions (HR 4.04, 95% CI 1.63-10.03) and not mixed/active inactive lesions (HR 0.89, 95% CI 1.88-3.95). Conversely, an increasing proportion of inactive lesions was associated with a slower time to death (HR 0.19, 95% CI 0.36-0.69) without impacting time to wheelchair (HR 0.99, 95% CI 0.52-1.90) (**Fig. 8i-j**).

**Fig. 8.**
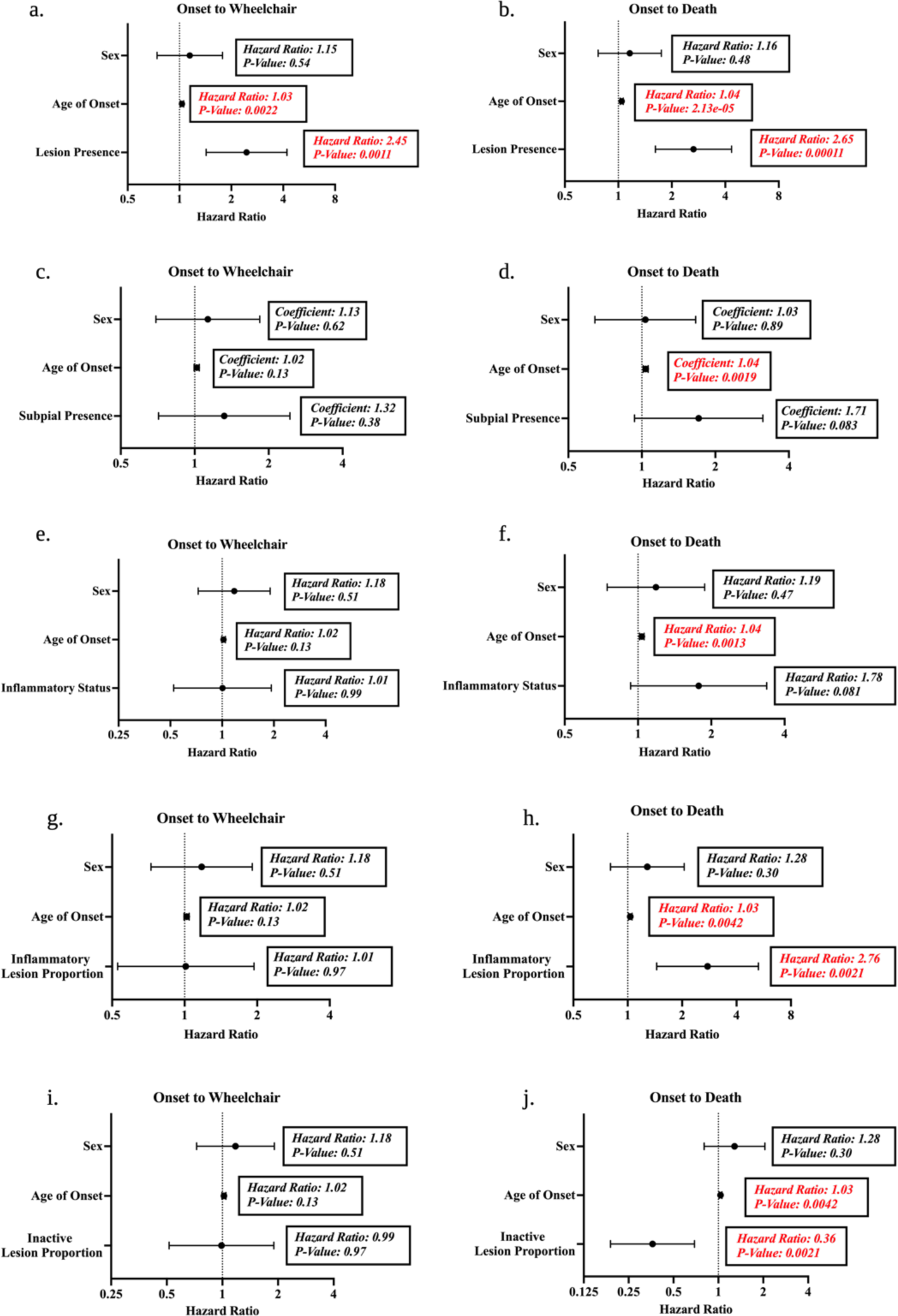
Clinicopathological Relationships Between Demyelination and Clinical Milestones. Forest plots displaying the hazard ratio and 95% confidence interval are derived from multivariate cox proportional hazards models controlling for sex and age of onset. Both case-level (a-f) and lesion-level (g-j) analyses were performed. The impact of spinal cord lesion presence on the time from MS onset to wheelchair use (**a**) and MS onset to death (**b**) was tested. In cases with lesions, the impact of subpial lesion presence was tested on the time from MS onset to wheelchair use (**c**) and MS onset to death (**d**). The impact of inflammatory status was also tested on the time from MS onset to wheelchair use (**e**) and MS onset to death (**f**). Inflammatory and inactive lesion proportions were tested on the time from MS onset to wheelchair use (g,i) and MS onset to death (h,j) Coefficients and p-values are highlighted in red for significant predictors (p<0.05) in the models.

## Discussion

Demyelination is a cardinal feature of MS spinal cord pathology and its topology has been of great interest. However, different studies have yielded discrepant interpretations and engendered considerable debate [16, 32, 33]. In our dedicated study of MS spinal cord lesional pathology, we find that inflammatory lesions in the MS spinal cord are the most prevalent, contrasting the traditionally held view that spinal cord lesions are ‘burnt out’ at end-stage disease. Further, our topographic finding that lesions most commonly affect the posterior and lateral columns with relative sparing of the subpial and central canal interfaces in a distribution that mirrors the vasculature suggests that an ‘outside-in’ gradient of diffusible CSF factors is not the sole culprit of lesion pathogenesis.

We found that lesions are commonly found at different spinal cord levels and are mostly inflammatory even in end-stage progressive MS. Previous MRI work has demonstrated a similar prevalence of spinal cord involvement to our study [7, 35, 44]. However, our study provides novel insight into the extent of inflammatory activity in spinal cord lesions at end-stage disease. Our finding of the high prevalence of lesional activity in spinal cord is in line with findings of independent autopsy studies placing an emphasis on supratentorial regions and the optic pathway [29, 34]. While MRI studies show that contrast enhancing lesions decrease over the MS lifecycle, it is well established that detection of active lesions, let alone mixed active/inactive ones, in the spinal cord is challenging [1, 12, 27]. The small size of the spinal cord, its proximity to mobile tissues and fluids, and relative shielding by bone and fat all contribute to the reduced sensitivity and specificity of lesion detection by MRI when compared to histopathology [9]. The fact that the significant majority of lesions at death remain inflammatory, particularly of the mixed active/inactive moiety, provides a new perspective on the traditional view that lesions become ‘burnt-out’ in the progressive phase and may not be amenable to anti-inflammatory therapy [18, 19].

We observed a modest preponderance of cervical lesions and the lesion status of the cervical spinal cord demonstrated high sensitivity in the prediction of lesions in the thoracolumbar region. Previous MRI studies in clinical cohorts also found spinal cord lesions to be more common in the cervical cord, but may have under-estimated thoracolumbar disease [7, 35, 44]. While technical and physiological artifacts reduce the sensitivity of lesion detection *in vivo*, the fact that we studied post-mortem specimens where a more significant accumulation of lesion burden is expected could also explain this difference. What is more, we found that the lack of cervical cord lesions predicted the absence of lesions in the thoracolumbar region. This data suggests that solely imaging the cervical spinal cord may provide important insights into the status of the thoracolumbar region. Therefore, particularly in resource-limited settings, prognostication and monitoring paradigms using spinal cord MRI could focus on just obtaining images from the cervical level.

We provide novel insight into the distribution and extent of demyelination in the MS spinal cord. In line with previous work, we found that MS cases most commonly have lesions affecting the posterior and lateral white matter columns and grey matter as a whole [14]. While the majority of MS cases harboured a subpial lesion (76/91 of cases, 83.5%), the proportion of the subpial surface affected by lesions was low (∼30% of cord circumference). Distance-based lesion segmentation analyses further supported the relative sparing of the subpial surface by showing that lesions most commonly affected the cord at distances furthest away from CSF boundaries (i.e. subpial and central canalicular surfaces). These findings differ from a recent MRI study that reported a subpial gradient of demyelination in the spinal cord [33]. Several factors could explain these divergent findings. Pathological studies have superior resolution to detect the presence and distribution of lesions, especially at CSF interfaces where radiographic signal-to-noise ratio is impaired. MRI studies of the spinal cord are further challenged by the cord’s proximity to mobile tissues and fluids and relative shielding by bone and fat [9, 30]. Hence, subpial signal changes previously reported on MRI may reflect pathological processes apart from demyelination. Reactive gliosis is a prime candidate as a subpial gradient of glial cell reactivity has been observed in the post-mortem spinal cord [22].

Our topographical analyses lend support to a role for the vasculature in the pathogenesis of demyelination in the disease. Collectively, our lesion probability maps and distance-segmentation analyses show that lesions mirror the vascular network with relative sparing of regions juxtaposed to CSF boundaries [16, 32]. We concluded that vascular pathology (e.g. blood-brain-barrier (BBB) disruption) is a likely contributor to the pathogenesis of the demyelinated lesion and argues against a primary ‘outside-in’ process of demyelination. In fact, while work in the cortex has related tertiary lymphoid follicles to cortical demyelination from the subpial surface inward, similar findings were not found in the spinal cord [36]. We also found that demyelination is underrepresented in the anterior funiculus, an area where CSF stasis and meningeal inflammation is posited to be at its highest. Further, cases with motor cortical subpial lesions did not have a greater extent of subpial demyelination in the spinal cord compared to cases without motor cortical subpial lesions. These findings suggest the relative importance of CSF-derived factors and the vascular unit may differ between the brain and spinal cord due to inherent topographical differences in 1) the mechanical forces imparted on these structures, 2) the intrinsic permeability of the BBB compared to the blood-spinal cord barrier (BSCB) [4, 32], and/or 3) dysfunction of CSF flow and clearance along perivascular spaces. However, it is plausible that the perivascular unit and neurotoxic CSF act in concert to promote lesion formation and sustain its inflammatory activity. Subpial cortical lesions are typically juxtaposed to areas of CSF flow but also within areas of vascularity [23]. In fact, the cortex is known to harbour a vascular-rich angiome [5]. Overall, a neurotoxic CSF may potentially exert its impact in areas of BBB/BSCB disruption via its flow within the perivascular spaces in addition to diffusion across a disrupted glia limitans into the parenchyma.

Indexed clinical reports with metrics of disease outcome obtained during life allowed us to draw interesting clinicopathologic correlations. The presence of a spinal cord lesion was an independent predictor of two important clinical endpoints, time from onset to wheelchair and onset to death. These findings confirm the relevance of spinal cord demyelination to MS prognosis purported by MRI studies of clinical cohorts [3, 8, 41]. In contrast to work from the Netherlands Brain Bank [29], cases with inflammatory lesions did not have a significantly worse clinical course compared to those with only inactive ones, a finding potentially explained by the small number of inactive cases in our cohort (n = 11). However, when performing a lesion-level analysis, the propoprtion of active lesions predicted a faster time from disease onset to death while the proportion of inactive lesions was a significant predictor of a longer disease duration. These findings are in line with recent post-mortem work using samples from the UK MS Tissue Bank that underscored the importance of supratentorial active lesions and perivascular inflammation in predicting a more severe disease course during life [31].

We acknowledge there are inherent limitations in post-mortem studies. Our cohort is necessarily biased towards end-stage MS. To address this, we included a large number of cases across a wide spectrum of clinical disability. Our focus on the post-mortem spinal cord sampled throughout its length is unique and provides resolution to detect granular cord pathology well beyond that of *in vivo* imaging modalities. This strategy enabled us to not only characterise lesion location more precisely but also to decipher the inflammatory activity of lesions and their distribution. While the study of three cord levels is comprehensive, the lack of inclusion of the sacral cord precludes conclusions specific to that cord level. Given that Wallerian degeneration and ischemia may lead to myelin changes, we defined primary demyelination with stringent criteria of complete myelin and absence of confounding pathology for inclusion in downsteam analyses. While there are limitations in the use of a single-marker (CD68) for lesion staging, we thought it important to implement the consensus lesion staging criteria for histological classification of MS lesions using myeloid markers for comparability across pathological studies. We recognise that our lesion maps represent a static view of a dynamic process and are an extrapolation from individual spinal cord sections that were deemed representative of the cervical, thoracic, and lumbar levels. However, we attempted to mitigate this by transposing lesions for each case into a standard space. While measurement error is possible, anatomical landmarks were used to maintain proportional relationships required for downstream analyses and reliability was assessed with two expert raters. Paired statistical analyses also improved statistical power. In comparison to dedicated spinal cord MRI, our approach did not allow for the comprehensive profiling of lesion topography throughout the entirety of the spinal cord. However, we placed emphasis on relevant heterogeneity within each section and across levels more broadly (cervical, thoracic, and lumbar). Overall, compared to MRI, our histopathological approach provides increased sensitivity and specificity for lesion detection and enhanced resolution for the assessment of inflammatory activity.

In summary, this study demonstrates that spinal cord lesions are an important feature of MS pathology. Spinal cord lesions are common, often inflammatory at end-stage disease, portend a worse clinical course, and exhibit a unique topographical pattern. The subpial zone is relatively spared across the cohort and lesion patterns mirror the trajectories of the vasculature (**Fig. 11**). These findings begin to disentangle the complexities surrounding the relative contributions of the meningeal interface and vascular unit to the pathogenesis of lesions in MS. Our study sets the stage for further exploration of the mechanisms that underlie topographical variation in lesion biology along the length of the MS neuraxis.

## Data Availability

The data that support the findings described in this manuscript are available from the corresponding author, GCD, upon reasonable request and execution of a data transfer agreement.

## Data Availability

All data produced in the present study are available upon reasonable request to the authors

## Acknowledgements

ADW was supported by the Emory University MD/PhD Program, NIH MD/PhD Partnerships Program, and NIH Oxford–Cambridge Scholars Program. MP was supported by the Oxford-Quinnipiac Partnership. MJ was supported by the NIHR Oxford Biomedical Research Centre (BRC), the Wellcome Trust, and Wellcome Centre for Integrative Neuroimaging. MJL was supported by the Division of Intramural Research, National Institute of Allergy and Infectious Diseases, NIH. GCD was supported by the NIHR Biomedical Research Centre (BRC), Oxford and has received research funding from the UK MS Society and the Oxford-Quinnipiac Partnership to support this work. The authors would like to thank the United Kingdom MS Tissue Bank as well as patient donors and their family members for the provision of spinal cord tissue and clinical information required to undertake this study. The authors also thank Margaret M. Esiri for her neuropathological expertise and guidance. The authors also acknowledge Jon Fitzi from the NIAID Biostatistics Research Branch and the GitHub community for biostatistical advice. Lastly, Fig. panels were created with BioRender.com using the National Institutes of Allergy and Infectious Diseases (NIAID) institutional license.

## Tables

**Supplementary Table 1.** Antibodies and Conditions Used for Immunohistochemistry.

| **Antibody** | **Clone** | **Classification** | **Antigen Retrieval** | **Dilution** | **Block** | **Incubation** | **Supplier** | **Catalogue** |
| --- | --- | --- | --- | --- | --- | --- | --- | --- |
| PLP | plpc | Mouse Monoclonal | Microwave, Citrate pH 6 | 1:1000 (TBST) | None | 1hr RT | BioRad | MCA839G |
| CD68 | PGM1 | Mouse Monoclonal | Microwave, Citrate pH 6 | 1:100 (TBST) | None | 1hr RT | DAKO | M0876 |

PLP=proteolipid protein, TBST=tris-buffered saline, 0.05% triton-X, RT=room temperature

## Figures

**Supplementary Fig. 1.**
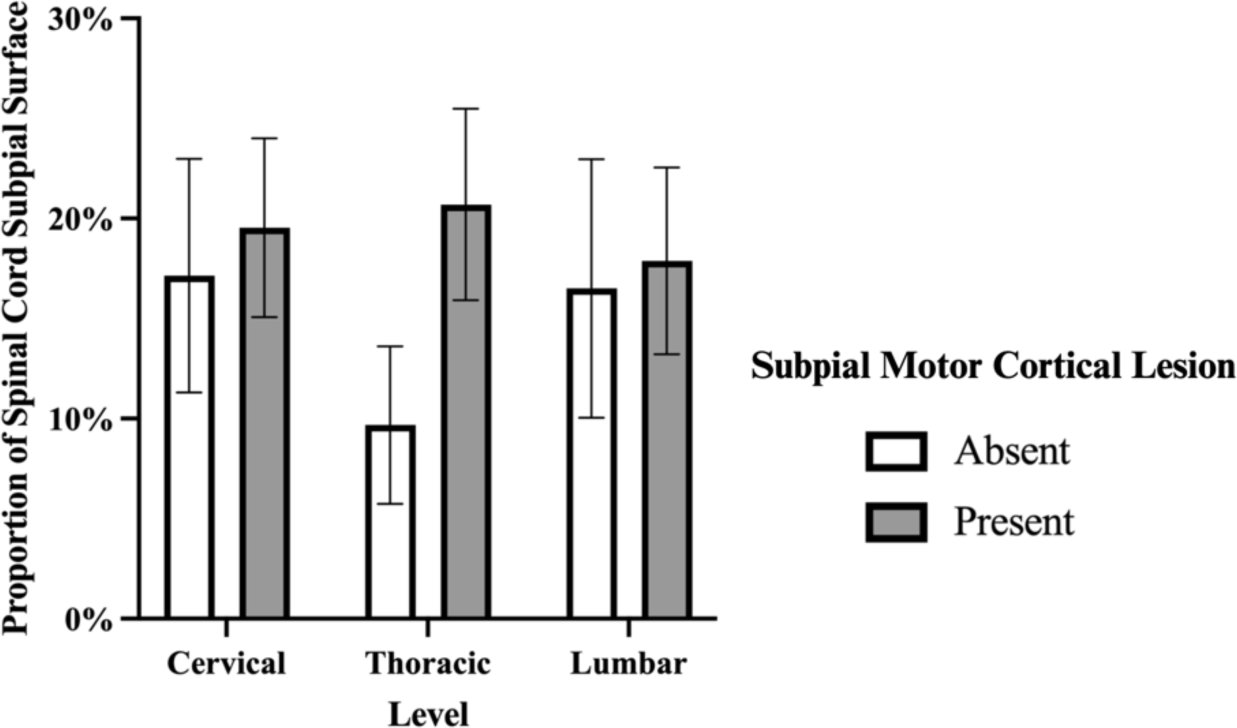
Relationship Between Subpial Demyelination Across Anatomical Sites. Grouped bar charts depicting the proportion of the spinal cord circumference impacted by subpial demyelination across levels in cases with subpial motor cortical lesions and those without. Proportions represent observed values, and the asterisks indicate significant post-hoc pairwise comparisons following quasibinomial mixed modelling and multivariate adjustment for multiple comparisons (*p<0.05; **p<0.01; ***p<0.001; ****p<0.0001).

**Supplementary Fig. 2.**
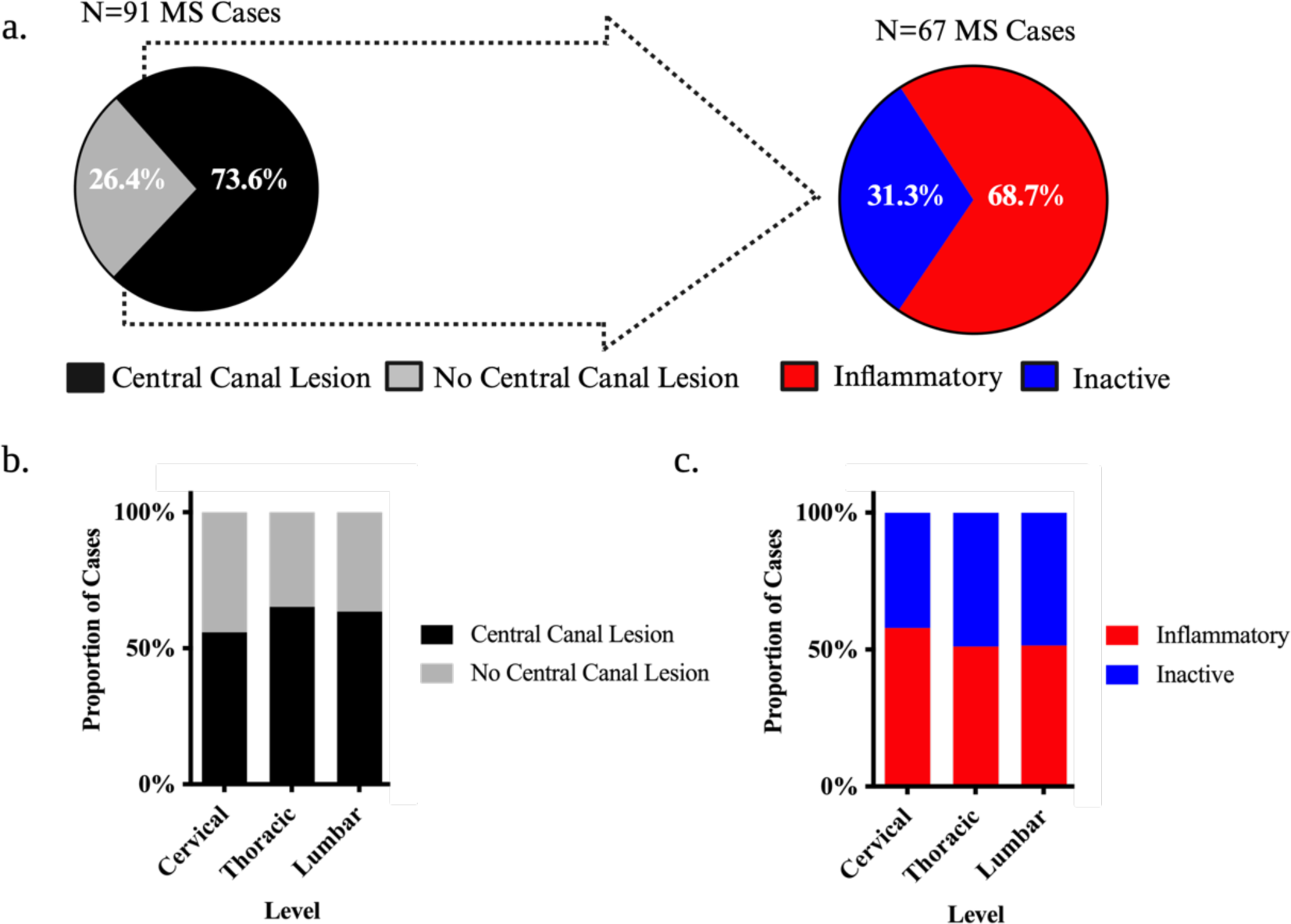
Prevalence of Demyelination in the Central Canal of the MS Spinal Cord. **(a)** Pie charts highlighting the observed proportion of cases with central canal lesions and those that harboured at least 1 inflammatory (active or mixed/active inactive) central canal lesion. (**b-c**) Stacked bar charts depicting the proportion of MS cases that harboured central canal lesions at each level of the spinal cord irrespective of stage (**b**) and classified by the presence of inflammation (**c**). Proportions represent observed values, and the asterisks indicate significant post-hoc pairwise comparisons following logistic mixed modelling and multivariate adjustment for multiple comparisons (*p<0.05; **p<0.01; ***p<0.001; ****p<0.0001).

